# Changes in Spatial Access to Obstetric Care in Germany from 2014 to 2024: A Nationwide Travel Time Analysis

**DOI:** 10.64898/2026.01.29.26345103

**Authors:** Maxi S. Kniffka, Niklas Ullrich-Kniffka, Loes C.M. Bertens, Jasper V. Been, D. Susie Lee, Josefine Königbauer, Amira Göpfrich, Jonas Schöley

## Abstract

**Background:** Timely access to healthcare is vital especially during childbirth, as it affects unplanned out-of-hospital births, survival and morbidity outcomes. In Germany, the number of maternity wards decreased since 2014, potentially increasing travel times for pregnant women. We examined changes in the travel times after maternity ward closures from 2014 to 2024 and addressed spatial disparities, providing essential information to ensure maternal and newborn care accessibility.

**Methods:** Maternity ward closures in Germany from 2014 to 2024 were identified, and travel time to the nearest ward was calculated for women of reproductive age (15 to 49 years) using the Open Source Routing Machine. Critical driving time was defined as 40 minutes or longer.

**Results:** Since 2014, the number of maternity wards in Germany decreased by 172 (-23.1%), leaving 573 in 2024. Consequently, the number of women facing critical travel times increased by 109%, from 48,139 (0.28% of all women at risk) to 101,025 (0.60%). 27 closures were responsible for 90% of the increase in critical travel times, with seven accounting for over 50%. Northern and eastern parts of Germany were affected most which was reflected in an increasing Gini coefficient measuring the inequalities in travel times across Germany.

**Conclusion:** Most maternity ward closures had minimal effect, but a few substantially increased travel time, especially in regions without nearby alternatives. These closures exacerbated regional disparities and potentially increased the risks of unplanned out-of-hospital births and other adverse birth outcomes.

**Key Messages:** *What is already known on this topic:* Long travel time to the nearest maternity ward is associated with unplanned out-of-hospital births and negative maternal, fetal and neonatal birth outcomes.

*What this study adds:* From 2014 to 2024, the number of maternity wards in Germany decreased by 23.1%. As a result the number of women between 15 and 49 facing travel times of 40 minutes or more increased by 109%, reaching 101,025 in 2024. Regional disparities in critical travel times were exacerbated.

*How this study might affect research, practice or policy:* This study underscores that further decisions on maternity ward closures should assess the impact on travel times and policy responses should create accessible accommodation options in areas where travel distance cannot be otherwise reduced. Further studies are needed to monitor the impact on increased travel time to obstetric care in Germany.

## Introduction

Timely access to healthcare services and hospitals plays a crucial role in ensuring population health, particularly during emergencies where swift access and short travel distances can be of major importance for survival [1–3] and morbidity outcomes [4].

While many healthcare needs can be planned and managed in advance effectively, the process of going into labor and giving birth constitutes a unique and delicate situation. In cases such as scheduled cesarean deliveries, the ability to plan ahead can mitigate the urgency of having a nearby maternity ward. However, in a country where the majority of women experience vaginal deliveries such as Germany [5], the timely availability of a maternity ward within a reasonable distance is crucial. This availability affects not only the safety of the mother and newborn but also the emotional and psychological comfort of the family. Long travel times have been linked to increased risks of fetal distress [6] and adverse birth outcomes, including perinatal mortality [7–9]. They are associated with up to a 2.5-fold increase in the risk of an unplanned out-of-hospital births [6,8,10], which independently increases the risk of perinatal mortality [7,11]. Unplanned out-of-hospital births are more common for young and multiparous women [7]. Extended travel distances can also elevate the risk of maternal morbidity [4]. Additionally, negative birth experiences can reduce women’s fertility intentions [12] and result in fewer subsequent children [13].

Definitions of long travel times vary across studies, ranging from over 20 minutes in the Netherlands [9] to more than 1 hour in Norway [8] based on their results. In Germany, the governmental committee “Gemeinsame Bundesausschuss” considers travel times above 40 minutes for gynecologic and obstetric care to be critical and recommends avoiding them [14].

However, travel time is not the only important factor when it comes to safe childbirth. Recent studies indicate that higher-level or higher-volume maternity wards are associated with lower maternal and perinatal mortality and morbidity, particularly for high-risk pregnancies and preterm-born infants [15–17]. For low-risk pregnancies, outcome differences between hospital levels and volumes are small, though slightly higher perinatal mortality has been reported in low-volume units [18]. These findings support a centralisation of high-risk obstetric care. Therefore, both the level of obstetric care and timely accessibility to appropriate maternity services are critical determinants of maternal, fetal and neonatal outcomes.

Due to centralisation efforts of obstetrics, several high-income countries show a reduction in the number of maternity wards. Following such closures, there has been an increase in out-of-hospital births by up to 200% in Australia [19], Finland [11], and Norway [20]. However, other studies from France [10], Norway [21] and Canada [22,23] indicate no negative effects of hospital closures.

Due to factors such as lack of perinatal expertise and resources for high-risk pregnancies [14], subsequent profitability, staff shortages or decreasing number of births [24], the number of maternity wards in Germany decreased from 865 in 2007 to 672 in 2017 [25]. These closures are likely to have affected travel times to the nearest maternity ward. Ward closures tend to disproportionately affect rural and socioeconomically disadvantaged regions, where recruitment of specialized staff is more challenging and birth volumes are lower [26]. This may exacerbate existing regional disparities in maternal and neonatal outcomes, particularly among populations with higher baseline obstetric risk [26–28].

This study examines the consequences of ward closures that occurred between 2014 and 2024 in Germany. We focus on how these closures influenced travel times to the nearest maternity ward for women in reproductive ages, considering spatial variations in small areas. Our goal is to quantify how travel times changed after these closures and address potential regional disparities. By highlighting this topic, we aim to contribute insights to healthcare policy and public health strategies, ensuring the accessibility of maternal and newborn care.

## Methods

Measuring the effect of maternity ward closures on spatial access in Germany requires high resolution geodata on the annual population at risk and the annual location of available maternity wards.

Women of reproductive age (15 to 49 years) were considered to be the population at risk, because they were, at least theoretically, ‘at risk’ of becoming pregnant. Data on the spatial distribution of women between 15 and 49 years was sourced from the UN Humanitarian Data Exchange, which provides high-resolution population estimates for Germany in 2019 [29]. This data was further aggregated into 1x1 km (0.62 x 0.62 miles) grids [30]. To account for population change over time, we used official annual county-level estimates of the number of women between 15 and 49 ages from 2014 to 2024 [31]. Using the spatial distribution data, we disaggregated the county-level population counts into 1×1 km grids, aligning the grids with the respective counties [32].

Information on maternity ward closures was derived from annual lists of all hospital births in Germany, publicly available from 2014 to 2024 [33]. This data included all maternity wards, detailing the number of births each year. If a ward reported births in one year but not in the next, we assumed it had closed. For wards where the number of births decreased by at least 15% or more compared to the previous year, we manually researched if and when it was closed. If a ward closed in November or December, the closure year was set as the subsequent year, because it had been available for most of the year. For years when a maternity ward did not report its birth numbers on time, but we confirmed it was still open, we used the previous year’s number of births to show the ward was active. The same was done if a maternity ward showed no births in one year, but at least one in the next year. In some cases a maternity ward moved locations within a bigger hospital campus or within a street. We did not consider those moves as a closure and a reopening of a new ward if the travel time between both addresses was less than 5 minutes by car and used the new address.

Travel time was calculated annually, representing the time required to drive from each 1x1 km grid to the nearest available maternity ward by car. For each grid we sampled 10 random points and calculated the mean driving distance. We used a locally hosted Open Source Routing Machine (OSRM), a routing service based on OpenStreetMap maps [34]. The map material comes from Geofabrik GmbH [35], which is an extract of the freely available OpenStreetMap maps [34].

Additionally, we calculated how many women in reproductive age were affected by critical driving times from each individual ward closure, by calculating the minimal driving time before and after each ward closure based on which wards were still available in each year. As these initial effects of ward closures are calculated with the population at risk from the year of the closure, the summed up effects of all closures do not completely add up to the overall increase in women at risk with critical travel time between 2014 and 2024. A critical driving time was set to 40 minutes or longer, following the official recommendation for Germany [14].

For state comparison we matched the grids and maternity wards to the belonging state [32] and aggregated the results by state. We calculated the annual population-weighted Gini-coefficient to evaluate changes in spatial inequality in driving times.

In Germany, every pregnant person is free to choose the hospital in which to give birth whereas high-risk pregnancies are typically referred to and managed in specialized expert centers [36]. However, the majority opt for the geographically closest facility [37], which is why we also only use the driving time to the closest ward for our analysis. We did not include any maternity wards in neighboring countries.

### Patient and Public Involvement

Patients or the public were not involved in the design, or conduct, or reporting, or dissemination plans of our research.

## Results

In 2014, there were 745 maternity wards providing delivery services in Germany. By 2024, this number had decreased by 172, resulting in 573 wards which equals a reduction of 23.1% (Figure 1). 173 wards were completely closed and 13 moved to a new location, resulting in 186 closures. One ward was newly opened. Although the majority of Germany had travel times of under 40 minutes in 2024 (indicated in green on Figure 2), there was a notable expansion of areas with travel times exceeding 40 minutes (indicated in purple).

**Figure 1:**
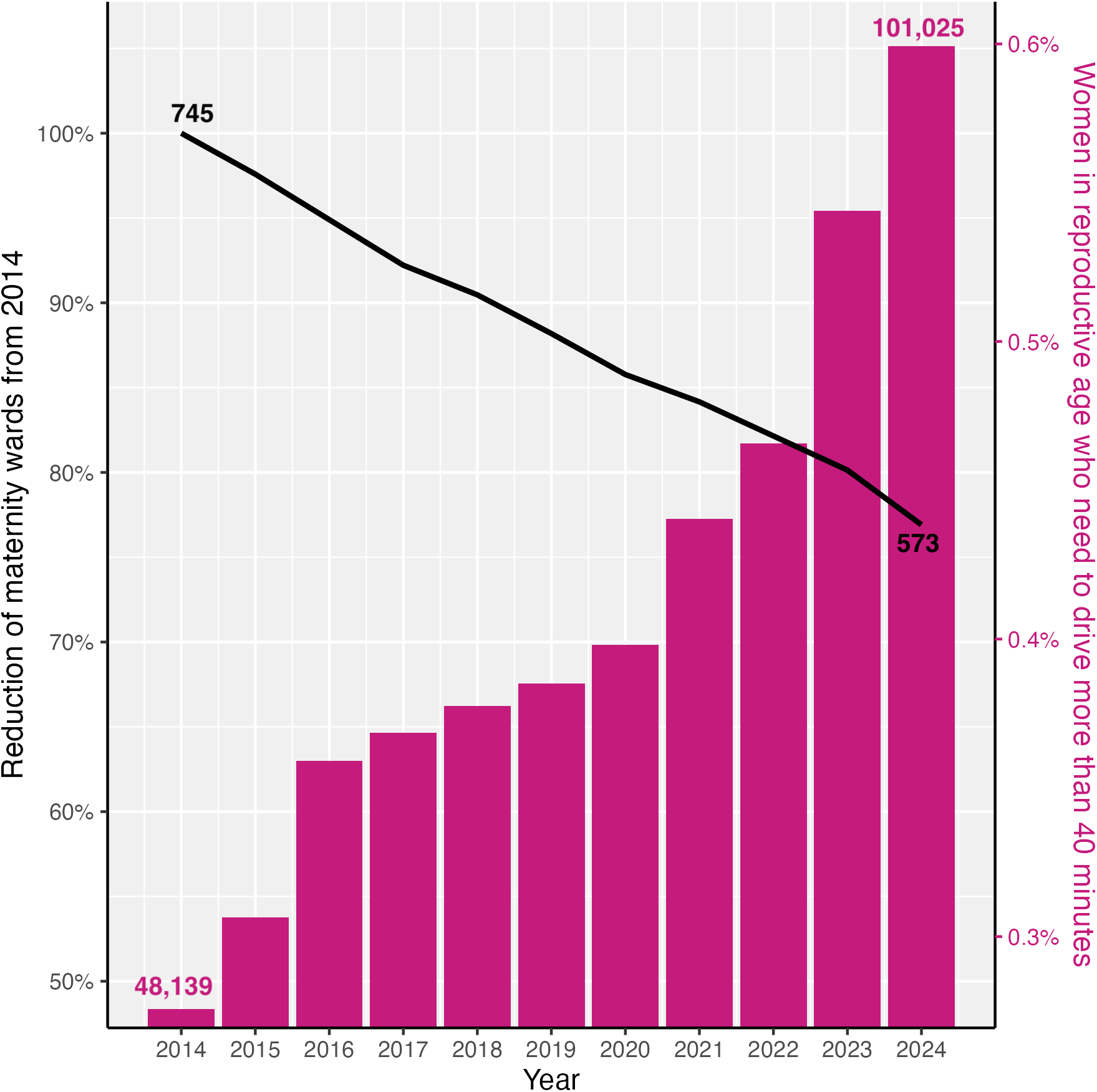
Change in available/ open maternity wards and the number of women at risk with critical driving times of over 40 minutes by car from 2014 to 2024 in Germany. Source: HDX (2019), Federal Statistical Office (2025) and Nutrica/ Milupa/ Danone (2014-2024). Calculated using OSRM.

**Figure 2:**
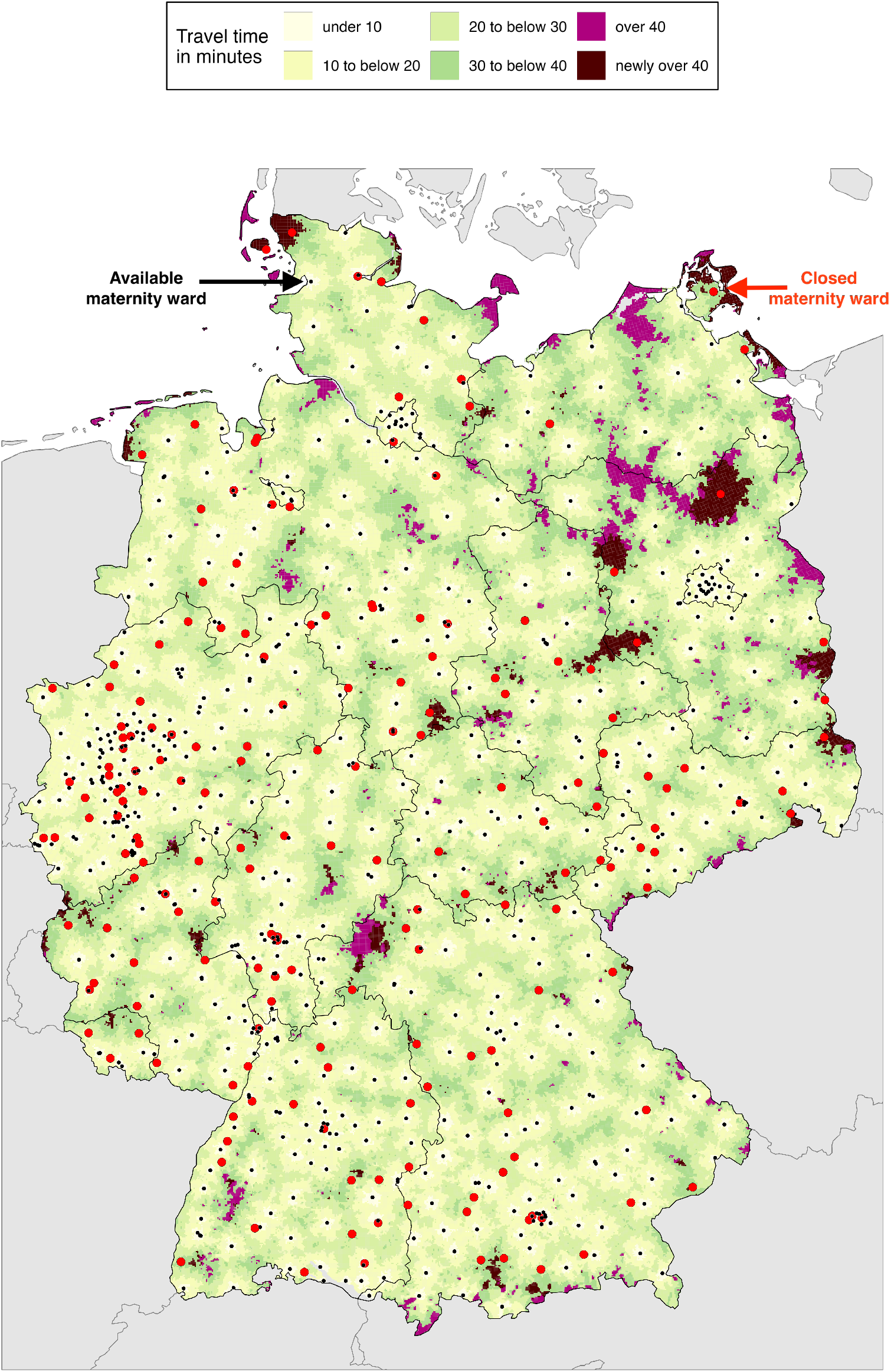
Travel time to the next available maternity ward in 2024 in minutes by car. Newly over 40 minutes refers to the change in travel time since 2014. Black dots mark available wards in 2024 and red dots represent wards that were closed between 2014 and 2024. Source: HDX (2019), Federal Statistical Office (2025) and Nutrica/ Milupa/ Danone (2014-2024). Calculated using OSRM.

From 2014 to 2024, the total number of women in reproductive age slightly decreased from 17,463,924 to 16,856,103 by 3.5%. At the same time, the number of women at risk with travel times of 40 minutes or longer increased by 109%, from 48,139 to 101,025 (0.28% to 0.60% of all women at risk). The number of women with critical driving time increased every year, rising more rapidly from 2021 onwards (Figure 1). The most affected areas were located along the northern coast and in eastern Germany (Figure 2). The Gini-coefficient was 0.303 in 2014, already showing moderate regional inequalities. Until 2024 it rose to 0.312 indicating an increase in spatial inequalities (Table 1).

**Table 1:** Descriptive findings from 2014 to 2024 in Germany. Source: HDX (2019), Federal Statistical Office (2025) and Nutrica/ Milupa/ Danone (2014-2024). Calculated using OSRM.

|  | 2014 | 2015 | 2016 | 2017 | 2018 | 2019 | 2020 | 2021 | 2022 | 2023 | 2024 |
| --- | --- | --- | --- | --- | --- | --- | --- | --- | --- | --- | --- |
| **Number of maternity wards** | 745 | 727 | 707 | 687 | 674 | 657 | 639 | 627 | 612 | 597 | 573 |
| **Women in reproductive age** | 17,463,924 | 17,436,464 | 17,279,347 | 17,132,115 | 16,994,565 | 16,864,874 | 16,721,106 | 16,643,649 | 16,694,843 | 16,795,237 | 16,856,103 |
| **With travel time in minutes to the next maternity wards of: N (%)** |  |  |  |  |  |  |  |  |  |  |  |
| **40 or more** | 48,139<br>(0.276) | 53,422<br>(0.306) | 62,031<br>(0.359) | 63,131<br>(0.368) | 64,141<br>(0.377) | 64,949<br>(0.385) | 66,571<br>(0.398) | 73,307<br>(0.440) | 77,765<br>(0.461) | 91,339<br>(0.544) | 101,025<br>(0.599) |
| **30 to under 40** | 329,045<br>(1.884) | 340,229<br>(1.951) | 355,477<br>(2.057) | 380,679<br>(2.222) | 383,613<br>(2.257) | 403,164<br>(2.391) | 423,280<br>(2.531) | 447,386<br>(2.688) | 477,144<br>(2.858) | 530,024<br>(3.156) | 582,664<br>(3.457) |
| **Under 30** | 17,086,740<br>(97.840) | 17,042,813<br>(97.742) | 16,861,839<br>(97.584) | 16,688,303<br>(97.409) | 16,546,812<br>(97.365) | 16,396,761<br>(97.224) | 16,231,254<br>(97.070) | 16,122,955<br>(96.872) | 16,139,934<br>(96.676) | 16,173,875<br>(96.300) | 16,172,414<br>(95.944) |
| **Gini-Coefficient** | 0.303 | 0.304 | 0.305 | 0.306 | 0.306 | 0.306 | 0.306 | 0.308 | 0.308 | 0.311 | 0.312 |

Out of the 186 maternity ward closures or moves, 27 were responsible for over 90% of the increase in women with critical travel times (Table 2). Seven of these closures alone accounted for over 50% of the rise in women with critical travel times. The remaining closures had a minor effect on spatial access to obstetric care.

**Table 2:** List of maternity ward closures responsible for over 90% of the increase in women at reproductive age with critical travel times of at least 40 minutes. Ordered by the number of new women at risk affected. Source: HDX (2019), Federal Statistical Office (2025) and Nutrica/ Milupa/ Danone (2014-2024). Calculated using OSRM.

| Location of closed maternity ward | Year of closure | New women at reproductive age with critical travel time: N (% of all new women) | Cumulative % of new women with critical driving time |
| --- | --- | --- | --- |
| Brandenburg | 2023 | 6,298 (10.9) | 10.9 |
| Mecklenburg-Western Pomerania | 2021 | 5,414 (9.4) | 20.3 |
| Brandenburg | 2024 | 4,243 (7.4) | 27.7 |
| Schleswig-Holstein | 2016 | 4,217 (7.3) | 35.0 |
| Mecklenburg-Western Pomerania | 2016 | 3,705 (6.4) | 41.5 |
| Saxony | 2023 | 2,588 (4.5) | 46.0 |
| Baden-Württemberg | 2022 | 2,585 (4.5) | 50.5 |
| Brandenburg | 2015 | 1,848 (3.4) | 53.8 |
| Brandenburg | 2020 | 1,861 (3.2) | 57.7 |
| Bavaria | 2023 | 1,850 (3.2) | 60.3 |
| Lower Saxony | 2024 | 1,707 (3.0) | 63.3 |
| Saarland | 2024 | 1,667 (2.9) | 66.1 |
| Bavaria | 2015 | 1,495 (2.6) | 68.7 |
| Saxony-Anhalt | 2018 | 1,371 (2.4) | 71.1 |
| Rhineland-Palatinate | 2019 | 1,365 (2.4) | 73.5 |
| Schleswig-Holstein | 2015 | 1,224 (2.1) | 75.6 |
| Rhineland-Palatinate | 2024 | 1,199 (2.1) | 77.7 |
| Rhineland-Palatinate | 2017 | 1,141 (2.0) | 79.7 |
| Lower Saxony | 2021 | 1,116 (1.9) | 81.6 |
| Brandenburg | 2023 | 902 (1.6) | 83.2 |
| Schleswig-Holstein | 2022 | 826 (1.4) | 84.6 |
| Rhineland-Palatinate | 2016 | 794 (1.4) | 86.0 |
| Hesse | 2023 | 593 (1.0) | 87.0 |
| Rhineland-Palatinate | 2015 | 562 (1.0) | 88.0 |
| Schleswig-Holstein | 2022 | 514 (0.9) | 88.9 |
| Bavaria | 2021 | 492 (0.9) | 89.8 |
| Saxony-Anhalt | 2022 | 488 (0.8) | 90.6 |
| All remaining maternity ward closures (N = 159) |  | 5,410 (9.4) | 100 |
| All new women at reproductive age with critical travel time |  | 57,574 |  |

The changes in maternity wards and the number of women with critical travel times vary between German states (Figure 3). The three urban city-states (Berlin, Bremen, and Hamburg) experienced almost no ward closures and saw no increase in critical travel times. In contrast, Brandenburg, Mecklenburg-Western Pomerania, and Schleswig-Holstein, located in northern and eastern Germany, experienced the biggest changes.

**Figure 3:**
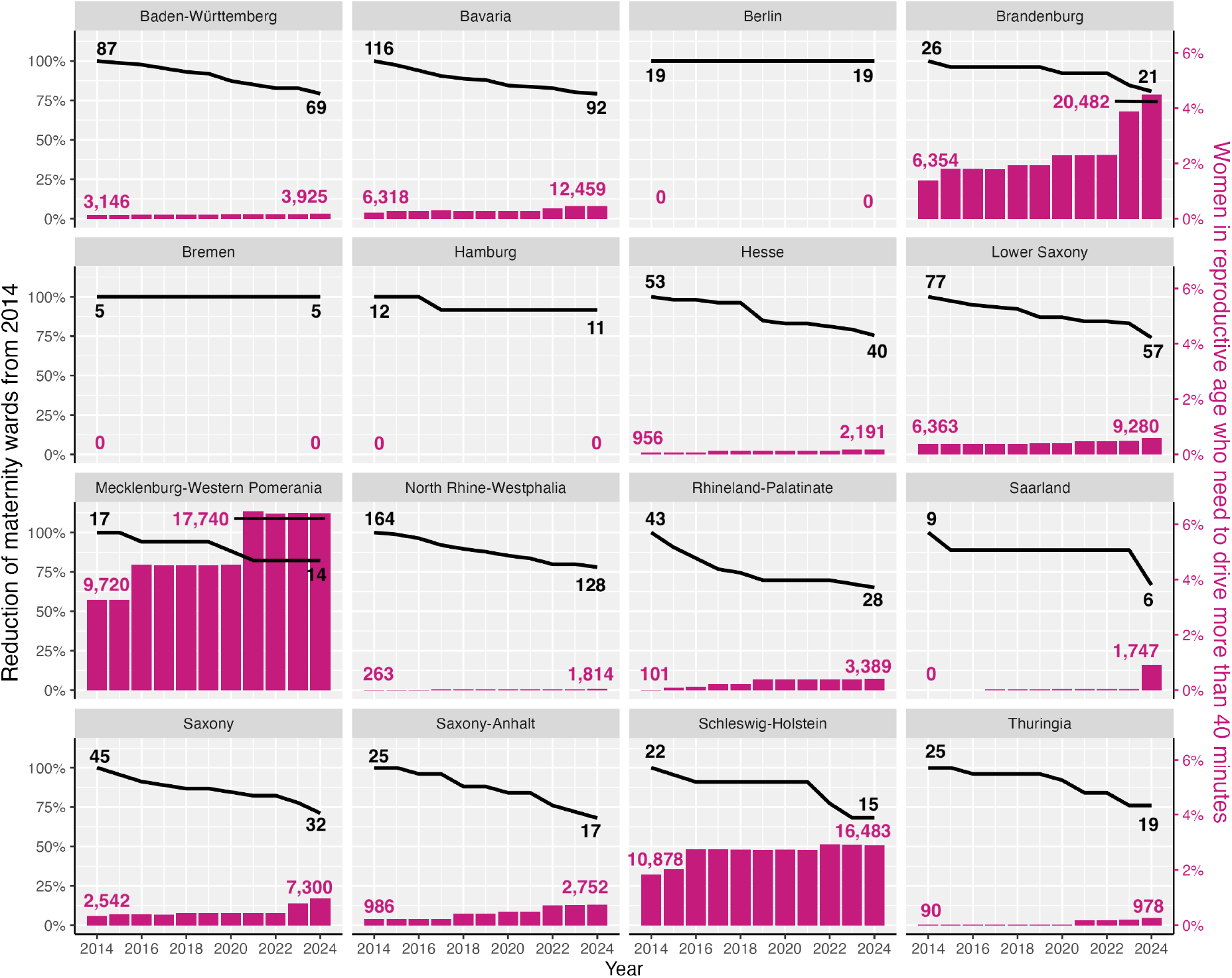
Change in available/ open maternity wards and women in reproductive age with critical driving times of at least 40 minutes from 2014 to 2024 by states in Germany. Source: HDX (2019), Federal Statistical Office (2025) and Nutrica/ Milupa/ Danone (2014-2024). Calculated using OSRM.

These states already had a high proportion of women living in areas with critical travel time in 2014, and they also saw the largest increases in such populations from 3.3% to 6.4% in Mecklenburg-Western Pomerania, 1.4% to 4.5% in Brandenburg and 1.8% to 2.9% in Schleswig-Holstein (Figure 3). Notably, 10 of the 27 maternity ward closures with the largest change in the critical travel times were located in these states (Table 2). After the closure of one maternity ward in Brandenburg, 6,298 women at risk newly faced travel times of more than 40 minutes, having previously been able to reach a maternity ward in under 40 minutes. This single closure accounted for 10.9% of the total increase in women with critical travel times. By comparison, the 159 ward closures that had the smallest impact on travel times together resulted in 5,410 new women at risk (9.4% of all newly affected women) needing to travel more than 40 minutes to reach the nearest maternity ward.

## Discussion

Between 2014 and 2024, the number of maternity wards in Germany declined by 23.1%. Over the same period, the number of women of reproductive age who must travel 40 minutes or more to reach the nearest maternity ward more than doubled, increasing by 109%. Most ward closures had little impact on geographical access to obstetric care. Rather, only 27 closures, representing just 15% of all closed or moved maternity wards, accounted for more than 90% of the increase in critically long travel times for women of reproductive age.

Several interrelated factors are driving the closure of maternity wards in Germany. Smaller units with low birth volumes often face unfavourable economics under the current hospital financing framework, where the costs of highly personnel-intensive services like midwifery and obstetrics are not adequately reimbursed. Simultaneously, persisting shortages of qualified midwives and physicians make it difficult for small facilities to maintain safe and sustainable services. Such dynamics have led to consolidation towards centres with higher case loads and specialist expertise, under the rationale of improving quality for high-risk pregnancies [14].

When maternity wards close in areas with alternative facilities nearby, spatial accessibility to obstetric care remains largely unaffected. However, closures of remote maternity wards result in substantially longer travel times to the next available facility. This can necessitate that women travel more than 40 or even 60 minutes while in labor, which has been shown to reduce the likelihood of facility-based delivery and negatively influence maternal, fetal and neonatal outcomes [4,6–11].

We find that the consequences of ward closures were not uniform across Germany. In 2024, Mecklenburg-Western Pomerania, Brandenburg and Schleswig-Holstein experienced notable increases in the proportion of women of reproductive age facing travel times of 40 minutes or more (to up to 6.5%). In these states, already high baseline travel times were exacerbated, entrenching regional inequalities in timely access to obstetric care. Such spatial disparities in access likely contribute to the heterogeneous findings in recent studies assessing the effects of maternity ward closures, since the surrounding hospital network and distances critically shape how closures translate into real-world access constraints [10,11,19–23].

Our study highlights the need to mitigate the effects of extended travel distances. Provision of incentives to work in remote units could support rural hospitals with difficulties finding qualified midwives and obstetricians. Another potential strategy is to ensure that expectant parents have access to temporary, accessible housing close to a maternity facility as they approach their due date. Evidence from Korea supports the concept of maternity waiting homes, which are residential facilities located near obstetric centres, as an intervention that reduces distance barriers and can improve maternal and neonatal outcomes. Research from obstetrically underserved areas in Korea found that women who used a maternity waiting home in the final weeks of pregnancy had shorter travel times to the maternity hospital and delivered infants with higher birth weights compared with women who did not use the facility, suggesting that proximity to a delivery facility can improve accessibility and perinatal outcomes [38]. The concept of maternity waiting homes or similar arrangements in order to reduce travel times might be especially important for women with second or third pregnancies as the risk for an unplanned out-of-hospital birth is highest for them [7], as labour might progress quickly.

Our study provides valuable insights in the current situation on spatial access to obstetrics in Germany. While the number of maternity wards reported here may differ slightly from official statistics due to data source limitations, the overall patterns of accessibility and regional disparities are robust, as they are in line with previous publications [24,25,27].

While this study focuses on spatial access to obstetric services, it does not assess the potential effects of increased travel times on maternal, fetal, and neonatal outcomes, which represents an important limitation. Unplanned out-of-hospital births have been reported to increase in several countries following a reduction in the number of maternity wards [11,19,20]. In Germany, it is currently not possible to assess whether a similar trend exists, as unplanned out-of-hospital births are not routinely monitored. Given the importance of evaluating the consequences of maternity ward closures, we highly recommend the systematic and routine surveillance of unplanned out-of-hospital births in Germany at state-level.

Closing maternity wards, or clinics in general, may be necessary in some cases, especially when workforce shortages make it impossible to guarantee high-quality, around-the-clock care. However, such closures must be carefully planned and coordinated to prevent unintended deterioration in care access. Future decisions on maternity ward closures should systematically assess the impact on travel times, and policy responses should include targeted support for maintaining or creating accessible accommodation options for expectant parents in areas where travel distance cannot be otherwise reduced. A small proportion of closures were responsible for the majority of increases in critical travel times in Germany, which underscores the importance of considering the geographic implications of closure decisions.

## Conclusion

While centralisation of obstetrics may improve quality, closure decisions must consider geographic access, ensure emergency readiness, and include targeted mitigation strategies to safeguard safe obstetric care.

## Declarations

### Ethical approval and consent to participate

This research project did not require ethics approval as it uses only publicly available data.

### Consent for publication

Not applicable

### Availability of data and materials

All data is available publicly. R-code is available on github: https://github.com/mskniffka/spacialchange

### Author contributions

MSK, NUK, and JS contributed to conceptualization. MSK contributed to data collection and curation, methodology and writing - original draft. MSK and NUK contributed to data analysis and visualization. MSK, NUK, JS, LCMB, JVB, DSL, JK and AG contributed to interpretation and writing - review and editing.

## Acknowledgements

We gratefully acknowledge the resources provided by the International Max Planck Research School for Population, Health and Data Science (IMPRS-PHDS).

